# DODGE: Automated point source bacterial outbreak detection using cumulative long term genomic surveillance

**DOI:** 10.1101/2024.01.21.24301506

**Authors:** Michael Payne, Dalong Hu, Qinning Wang, Geraldine Sullivan, Rikki M Graham, Irani U Rathnayake, Amy V Jennison, Vitali Sintchenko, Ruiting Lan

## Abstract

**Summary:** The reliable and timely recognition of outbreaks is a key component of public health surveillance for foodborne diseases. Whole genome sequencing (WGS) offers high resolution typing of foodborne bacterial pathogens and facilitates the accurate detection of outbreaks. This detection relies on grouping WGS data into clusters at an appropriate genetic threshold, however, methods and tools for selecting and adjusting such thresholds according to the required resolution of surveillance and epidemiological context are lacking. Here we present DODGE (Dynamic Outbreak Detection for Genomic Epidemiology), an algorithm to dynamically select and compare these genetic thresholds. DODGE can analyse expanding datasets over time and clusters that are predicted to correspond to outbreaks (or ‘investigation clusters’) can be named with the established genomic nomenclature systems to facilitate integrated analysis across jurisdictions. DODGE was tested in two real-world genomic surveillance datasets of different duration, two months from Australia and nine years from the UK. In both cases only a minority of isolates were identified as investigation clusters. Two known outbreaks in the UK dataset were detected by DODGE and were recognised at an earlier timepoint than the outbreaks were reported. These findings demonstrated the potential of the DODGE approach to improve the effectiveness and timeliness of genomic surveillance for foodborne diseases and the effectiveness of the algorithm developed.

**Availability and implementation:** DODGE is freely available at https://github.com/LanLab/dodge and can easily be installed using Conda.

**Supplementary information:** Supplementary Tables, Results, Figure 1 and Figure 2

## Introduction

Foodborne pathogens are a major cause of morbidity globally with 550 million infections reported in 2010 (Kirk *et al*. 2015). *Salmonella enterica* is a common cause of these infections with 78 million cases per year with the two most common serovars being *S*. Typhimurium (STM) and *S*. Enteritidis (Hendriksen *et al*. 2011, Kirk *et al*. 2015). Once they reach the human population from agricultural and environmental reservoirs the control of these pathogens depends on the identification and elimination of outbreaks. Outbreaks are mostly caused by single strains that contaminate food and lead to many cases of disease over a short time span. The identification of an outbreak has therefore relied on identifying strains that share the same genetic or phenotypic makeup and have occurred over a short temporal window (Sabat *et al*. 2013).

Whole genome sequencing (WGS) has offered new capacity to identify related clinical and food isolates at high resolution. Previous studies have demonstrated that isolates within an outbreak examined using WGS are often not genetically identical but are very closely related (Octavia *et al*. 2015). Therefore, there is a need to group isolates together using a genetic distance threshold. A single static genetic threshold is unlikely to be universally applicable due to differences in genetic diversity across bacterial populations and differences in the transmission pathways within the outbreak (Bekal *et al*. 2016, Gymoese *et al*. 2017, Leekitcharoenphon *et al*. 2014, Octavia *et al*. 2015, Phillips *et al*. 2016). We previously demonstrated the utility of a variable genetic threshold that depended on the local diversity of isolates over time and provided optimal sensitivity and specificity for outbreak detection (Payne *et al*. 2019).

Therefore, there is a need for a method and software tool that can identify an outbreak using thresholds determined dynamically based on the population and evolutionary dynamics of the pathogen. Public health genomic surveillance has become routine in many countries therefore any software solution must also be capable of identifying and tracking outbreaks over time in continuously expanding datasets.

In this study we present DODGE (Dynamic Outbreak Detection for Genomic Epidemiology), a method and tool to identify outbreaks with dynamic genetic thresholds selected using temporal thresholds that can accommodate expanding datasets from ongoing surveillance (software available from https://github.com/LanLab/dodge). This method utilises retrospective genomic surveillance data to define a background dataset which is then used to identify distinct, new clusters that subsequently appear. The method was tested on two STM genomic datasets, genome sequences from all STM isolates from a two-month period from two Australian states and over 9 years from the United Kingdom.

## Materials and Methods

### DODGE inputs

DODGE is primarily designed for use with cgMLST allele profiles and can accept this data directly downloaded from MGTdb or Enterobase (Kaur *et al*. 2022, Payne *et al*. 2020, Zhou *et al*. 2020). Temporal and nomenclature data (MGT STs or hierCC clusters) were extracted from metadata files that can be obtained from the corresponding databases. To facilitate ad hoc analyses using DODGE, SNP based inputs can also be used. Inputs using SNP analysis are vcf files and masked genomes produced by the program snippy (Seemann 2015).

### DODGE algorithm

In order to identify genetic clusters of bacteria that are likely to correspond to point source outbreaks DODGE uses a temporal threshold to dynamically select the best genetic threshold for each cluster independently. The stages of the DODGE algorithm are as follows (Figure 1A and B). Firstly, calculate pairwise distances between all isolates and perform single linkage clustering, clusters at each allowed thresholds are saved. Secondly, identify all genetic clusters at the maximum single linkage distance allowed (e.g., 5). For each cluster, if it is above the minimum size (e.g., 5 strains) check for the timespan of the collection times of isolates within the cluster. For clusters with timespans greater than the temporal threshold (e.g., 28 days) reduce the genetic threshold by one and check temporal threshold again. This is repeated until the timespan of the cluster is below the temporal threshold. This cluster is then stored as an investigation cluster which denotes a cluster that may warrant further examination using more detailed traditional epidemiological analysis. The minimum genetic threshold that retains the initial investigation cluster size is selected (e.g., if a cluster with threshold of 3 and 2 are identical threshold of 2 will be retained).

**Figure 1.**
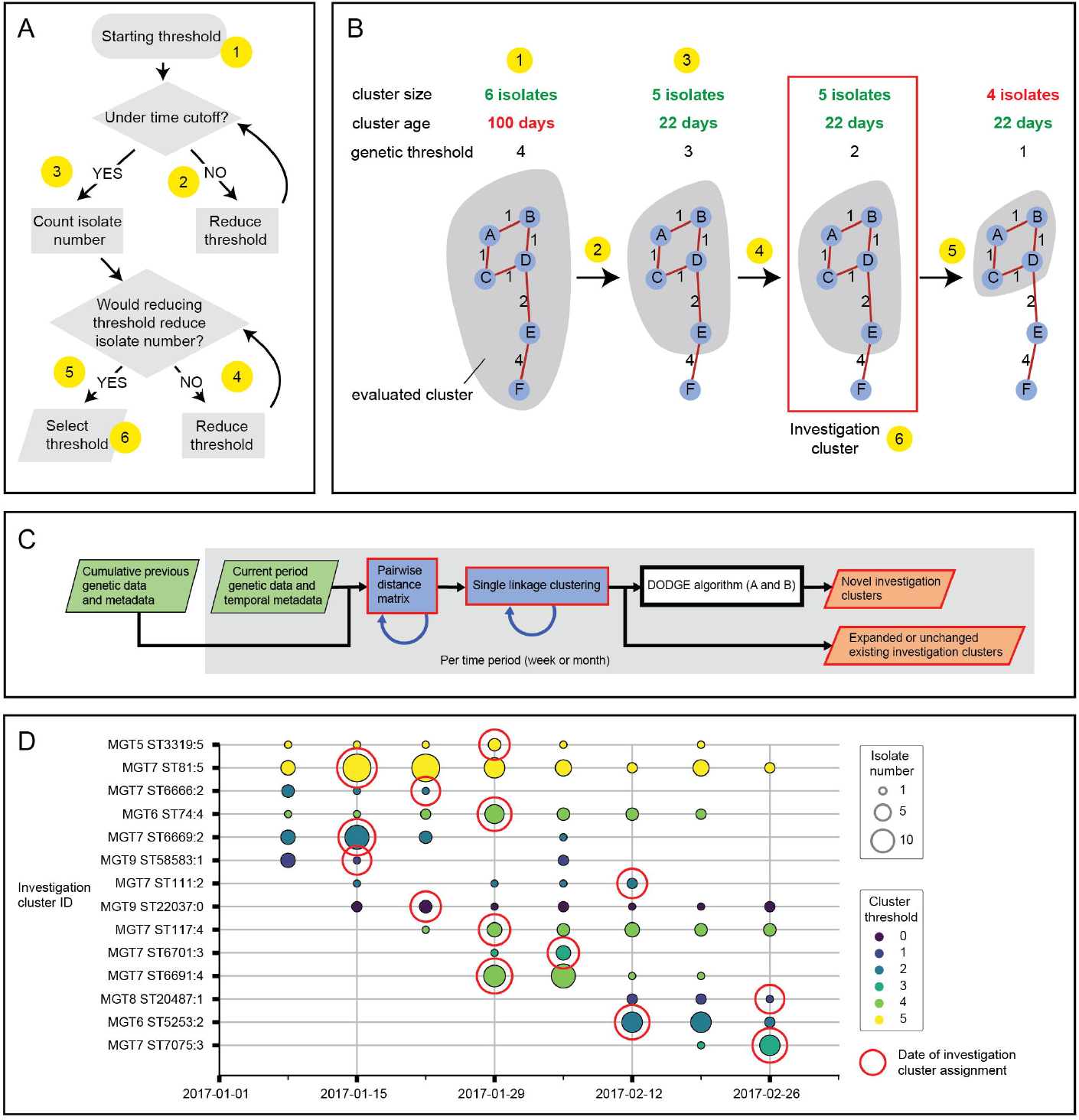
The DODGE pipeline, algorithm and Australian dataset investigation clusters. **A**. Flowchart describing 6 stages of the DODGE algorithm. **B**. Example investigation cluster detection with the same 6 stages marked. Blue circles represent isolates, red numbered lines are genetic distances. At each genetic threshold isolates within the grey shaded area are the cluster being evaluated. **C**. High level schematic of the DODGE pipeline including the DODGE algorithm. Genetic data in the form of allele profiles (Enterobase or MGTdb) or SNPs (output by snippy) for isolates from a given temporal window (a week or month) are combined with previous time periods to generate a combined distance matrix. Distances between isolate pairs that are not in an optional input distance matrix (Blue arrow) are calculated and added. Clusters are identified using single linkage clustering from the distance matrix. These clusters are compared to existing investigation and non-investigation clusters from previous time periods (blue arrow) to identify expanded or unchanged investigation clusters. Remaining non investigation clusters are then used to identify novel investigation clusters using the DODGE algorithm detailed in B and C. Green boxes are input files, red outlined boxes are output files, blue arrows represent outputs from one time period used as inputs in the next. **D**. Investigation clusters identified from the Australian dataset over time. X axis is date of collection by week. Y axis is investigation cluster with MGT ST based ID. The area of circles is proportional to number of isolates in that investigation cluster in that week. Colour represents the genetic threshold used for that investigation cluster. Red outline indicates the week in which the cluster was identified as an investigation cluster by the DODGE algorithm.

For the temporal window to be effective in selecting genetic thresholds a set of background isolate data should also be included. This background dataset is composed of isolates collected before the start date of the main investigation and is processed by DODGE to identify existing genetic clusters without calling any for investigation. Any cluster that originated in this time period is treated as background and will not be reported.

To track investigation clusters over time, each cluster is named based on the genomic identities of its constituent isolates and the genetic threshold used to identify it. For cgMLST data obtained from the MGTdb website each investigation cluster will be assigned an MGT ST. For data obtained from Enterobase a hierCC cluster name will be assigned (Zhou *et al*. 2021). The name is selected at the highest resolution level where greater than 70% of isolates in the cluster share the same ST (for MGT) or cluster (for hierCC). For example, in a cluster of 20 isolates, 20 (100%) have the same MGT6 ST, 17 (85%) the same MGT7 ST and 12 (60%) the same MGT8 ST, the MGT7 ST is then chosen as the investigation cluster name.

The same process is used for hierCC progressing from larger to smaller threshold hierCC clusters. Because SNP based analyses have no standardised nomenclature, numerical investigation cluster names are assigned per analysis. The second part of the investigation cluster name is the genetic threshold chosen by DODGE so that the full name is “genomic identity:genetic threshold”.

### DODGE pipeline

Most genomic analyses operate on a static set of isolates. However, pathogen surveillance occurs continuously, and the analyses must be able to absorb additional isolates on a regular basis. For this reason, the DODGE pipeline was designed to run the DODGE algorithm with any input dataset divided into segments (1 week or 1 month) and runs once for each segment (Figure 1C). For example, if a dataset contains 2 months of data and the time period was set to week based then the DODGE pipeline would run 1 background run on data sampled from prior to those 2 months and then 9 separate detection runs, one for each of the 9 weeks in the 2 months. The first run would identify clusters in the background dataset. Each subsequent detection run would include all previous investigation and non-investigation clusters (from previous weeks and background) and would identify if an investigation cluster was new, expanded or unchanged from one week to the next. In this way DODGE produces the same results from a large dataset over multiple years whether that data was added prospectively week by week or retrospectively in one run. Importantly investigation cluster names assigned by the DODGE algorithm are inherited across time periods to allow ongoing surveillance and tracking of the cluster. Additionally, once the cluster is identified as an investigation cluster, temporal thresholds used for cluster identification are no longer applied to allow long lived investigation clusters to be reported. The DODGE pipeline can also be run with a single static genetic threshold (bypassing the DODGE algorithm) if needed.

### DODGE case study datasets and algorithm settings

The Australian dataset includes genomic data for all STM isolates collected and sequenced at NSW and QLD public health laboratories in January and February 2017. All isolates in the STM MGTdb from Australia before 2017 were used as the background dataset. DODGE was run on this dataset with 5 isolate minimum cluster size and 28-day temporal threshold (ie, 5 cases in a 28 day window as signal of an outbreak) and an initial genetic threshold of five.

For the UK dataset all STM isolates from the UK from 2014 to 2022 that had year and month metadata were extracted from the STM MGTdb database. DODGE was run using a 5 isolate minimum cluster size, 5 genetic distance maximum and a two month temporal window.

## Results

### Application of DODGE to two months of Australian surveillance data using MGT

A total of 517 STM genomes from NSW and QLD sequenced in January and February 2017 were used to identify investigation clusters using DODGE (data available at https://github.com/LanLab/dodge/tree/main/examples/). Existing publicly available Australian isolates collected prior to 2017 were used as background data and included 1030 isolates over 26 years (Supplementary Table 1). Fourteen investigation clusters including 214 isolates (41.4%) were identified from the 2 months of surveillance (Figure 1D, Supplementary Figure 1, Supplementary Table 2). The average investigation cluster timespan was 29.3 days, average size was 15.3 isolates and average maximum pairwise distance was 4.7 allele differences. Of the 208 isolates in investigation clusters, 35 (16.8%) were collected before the cluster was identified as an investigation cluster, 77 (37.0%) were collected in the week the cluster was identified and 96 (46.2%) were collected after the identification. The Australian dataset was also run using SNP inputs and investigation clusters showed good agreement with MGT based clusters with a kohens kappa score of 0.91 (Supplementary Results).

### Application to UK data

Publicly available STM genomic surveillance data from the United Kingdom between 2014 and 2022 was evaluated as it is the most complete large dataset that includes month and year metadata (n=13251, Supplementary Table 3). Isolates from 2014 and 2015 were used as background data (n=2912) and the remaining 7 years of isolates (n=10339) were used to detect investigation clusters. A total of 93 investigation clusters were identified (Supplementary Table 4, Supplementary Figure 2) containing 1727 isolates (16.70% of 7 year dataset). The average investigation cluster timespan was 9.19 months, average size was 19.38 isolates and average maximum pairwise distance was 3.75 allele differences. Of the 1727 investigation cluster isolates, 105 (6.1%) were collected before the corresponding cluster was identified as an investigation cluster, 719 (41.5%) were collected in the week the corresponding cluster was identified and 909 (52.4%) were collected after the identification.

Two epidemiologically confirmed outbreaks were matched to publicly available representative genomic data within the UK dataset. The first was identified in April 2020 and caused 104 confirmed cases in the UK (European Centre for Disease Prevention and Control 2020). A representative from this cluster fell within the MGT9 ST22592:1 investigation cluster which contained 90 isolates and was assigned as an investigation cluster in February 2020.

The second outbreak was reported in February 2022 and consisted of two distinct clusters which caused 102 and 7 epidemiologically linked cases in the UK, respectively (Larkin *et al*. 2022). Two representatives for Cluster 1 fell within the investigation cluster MGT7 ST21164:4, which contained 108 isolates and was assigned as an investigation cluster in January 2022. Two representatives for Cluster 2 fell within investigation cluster MGT9 ST30910:3, which contained 7 isolates and was assigned as an investigation cluster March 2022.

## Discussion

The identification of point source outbreaks using automated methods often rely on epidemiological data such as time, location and strain phenotype without considering detailed genetic relationships between isolates (Latash *et al*. 2020, Salmon *et al*. 2016, Zhang *et al*. 2021). The increased uptake of WGS for prospective public health surveillance of different bacterial pathogens has the potential to provide this genetic context. However, the identification and reporting of emerging outbreaks from large datasets requires significant time and expertise. A recent promising approach for outbreak threshold detection employs temporal metadata and evolutionary modelling to select optimal genetic clusters (Duval *et al*. 2023). However, it was tested on a small set of simulated data with addition of only one real-life outbreak. Another published method does allow for clusters to be named and tracked over time from large datasets but does not select or adjust thresholds nor identify potential outbreak clusters (Mixao *et al*. 2023). A third method can identify whether an isolate should be included in an existing outbreak but cannot detect those outbreaks initially (Radomski *et al*. 2019).

DODGE is designed to identify a potential outbreak cluster by dynamically selecting the genetic threshold appropriate for the given investigation cluster using large, long term ongoing genomic surveillance datasets. This is achieved by identifying a genetic threshold for a given cluster that is stringent enough to exclude all isolates that occur more than a certain time in the past. In this way when sufficient background data is available, DODGE can adapt to the diversity of different clades in a population to provide more accurate outbreak cluster detection.

Other key features of DODGE are the ability to analyse data in on-going surveillance while maintaining cluster identity through existing bacterial genomic nomenclature systems (MGT and hierCC). These nomenclature systems have been applied to all public global data for STM allowing investigation clusters to be placed in broader genomic context while also facilitating simple communication of outbreak types.

Two investigation clusters from the UK dataset were matched with previously described outbreaks (European Centre for Disease Prevention and Control 2020; Larkin *et al*. 2022). In both cases representative isolates from the outbreaks were found within investigation clusters predicted by DODGE. These investigation clusters matched the size and timeframe reported for the outbreaks. Importantly, in both cases investigation clusters were identified prior to the date the cluster was originally reported (1 month earlier for MGT7 ST21164:4, 2 months earlier for MGT9 ST22592:1). This potential improvement in detection speed could allow more rapid responses to outbreaks, potentially reducing the overall number of outbreak cases. Additionally, in both the Australian and UK datasets, a significant proportion of isolates in investigation clusters were sampled after their respective investigation clusters were first detected (46.2% and 52.4%, respectively). These clusters represented likely community outbreaks and if they were investigated in a timely manner and preventative measures were implemented, the public health and societal burden of such clusters could be substantially reduced.

DODGE provides a means to identify, name and track outbreak clusters using dynamic thresholds from prospective genomic surveillance datasets and can be incorporated within laboratory surveillance and analysis workflows. The program is publicly available (https://github.com/LanLab/dodge) and could be used to accelerate the identification and control of point source outbreaks in any bacterial species where appropriate quality surveillance data is available.

## Supporting information

Supplementary Results

## Data Availability

All data produced in the present work are contained in the manuscript and at https://github.com/LanLab/dodge

## Acknowledgements

This work was supported by The National Health and Medical Research Council: [Grant Number 1146938].

