## Supplementary Figure 1 for "DODGE: Automated point source bacterial outbreak detection using cumulative long term genomic surveillance"

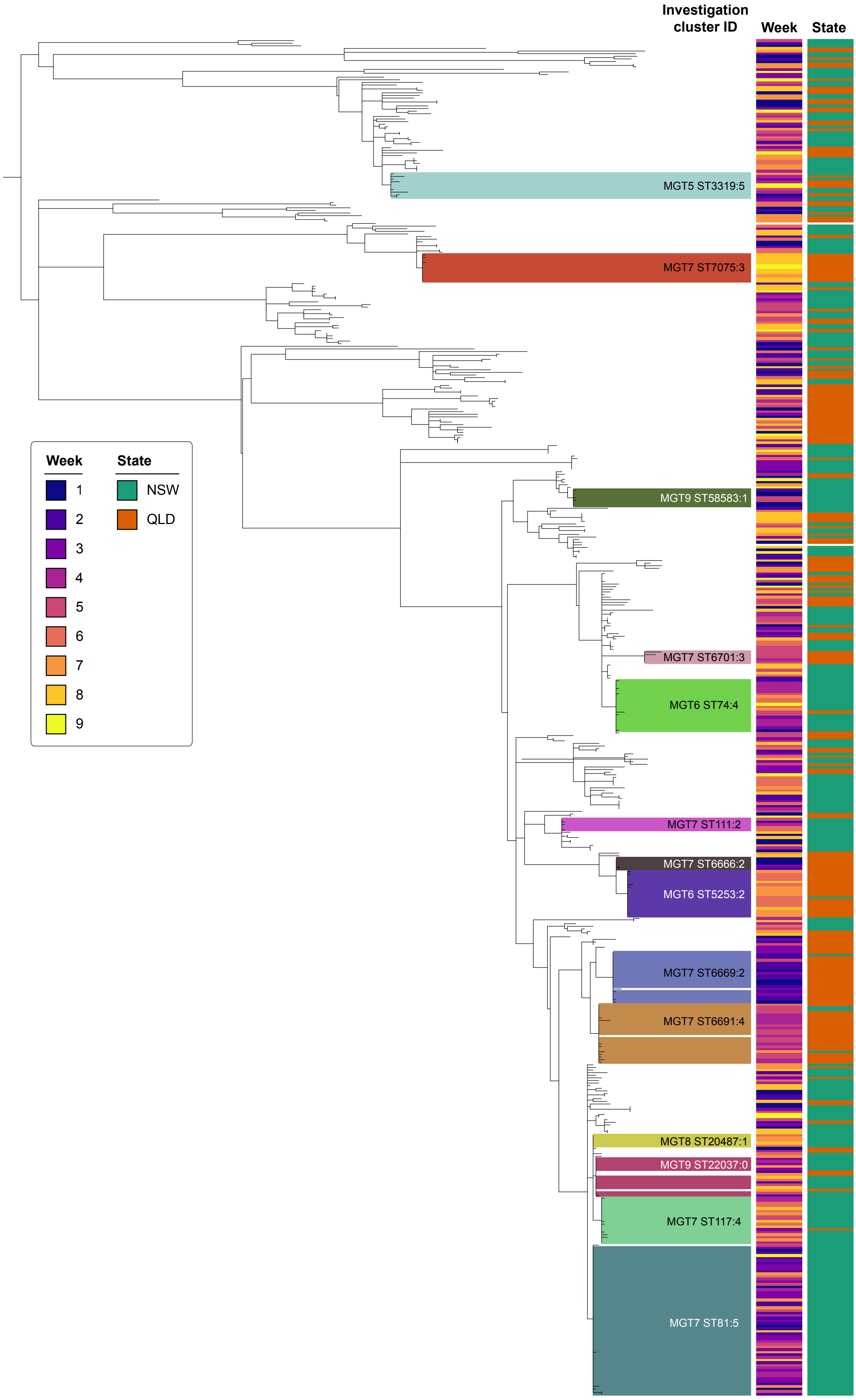


**Supplementart figure 1. Maximum likelyhood SNP tree of 2 month Australian data-set showing investigation clusters.** Investigation clusters are highlighted in different colours and their cluster ID is listed. Isolation week and state of each isolate is shown in corresponding colourstrips. Branches with ultrafast boostrap supports of less than 95 were removed.
