## Supplementary Figure 2 for "DODGE: Automated point source bacterial outbreak detection using cumulative long term genomic surveillance"

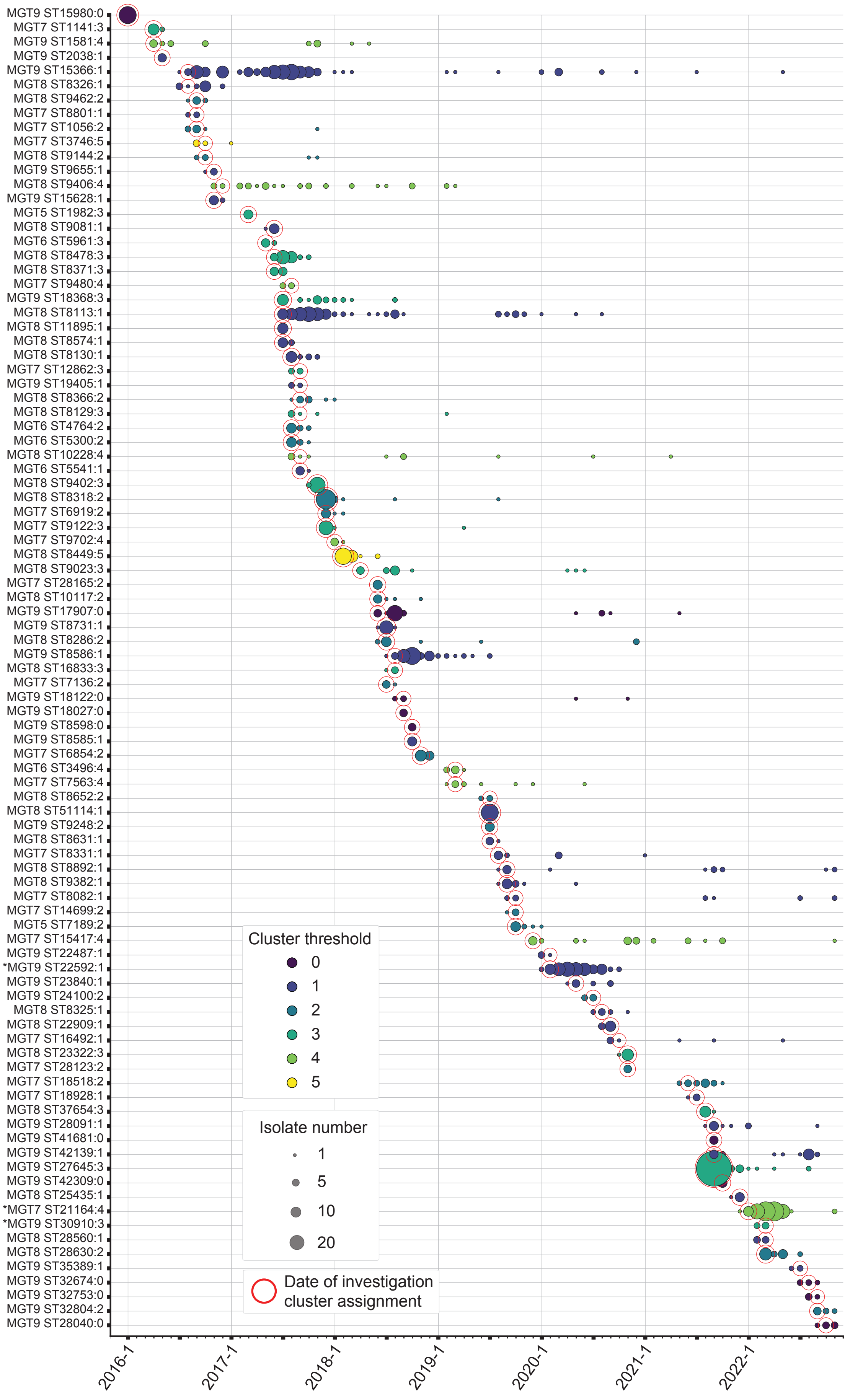


**Supplementary figure 2. Investigation clusters identified from the UK dataset over the 7 years examined.** X axis is month of collection. Y axis is investigation cluster with MGT ST based ID. The area of circles is proportional to number of isolates in that investigation cluster in that week. Colour represents the genetic threshold used for that investigation cluster. Red outline indicates the week in which the cluster was identified as an investigation cluster by the DODGE algorithm. * Indicates one of the clusters confirmed from published data
