## Supplementary Results for "DODGE: Automated point source bacterial outbreak detection using cumulative long term genomic surveillance"

**Relative performance of SNP and cgMLST data in the Australian dataset**

To evaluate the relative performance of SNPs and cgMLST alleles (from MGTdb) Australian dataset was also analysed using SNP based analyses. SNPs were identified from raw Illumina reads for all isolates using snippy version 4.6.0 with default settings. In the 2 month dataset SNPs have slightly higher resolution than MGT alleles due to the cgMLST scheme of MGT9 covering only 82% of the LT2 reference genome. Indeed 82.2% of SNPs were called in genomic regions included in the cgMLST scheme. Agreement of genetic variant assignment (alleles vs SNPs called in the corresponding locus) between cgMLST and SNPs was 94.7% with only 2.0% of variants unique to MGT and 3.3% of variants unique to snippy. Using SNP data, 11 investigation clusters with 212 isolates were identified. All 11 of these outbreaks were also identified when DODGE was run using MGT allele profiles. MGT identified 14 clusters in total. This three cluster discrepancy is composed of one cluster that was not detected using SNPs (id MGT5 ST3319:5, n=10) as well as three MGT based clusters that have merged to become one cluster in the SNP analysis (id MGT7 ST81:5, id MGT9 ST22037:0 and id MGT8 ST20487:1, n=72). Of the clusters that were detected by both methods (including the 3 that merged) 3 were identical, 2 were larger in the MGT analysis, 3 were larger in the SNP analysis and 3 had unique isolates from both methods. Overall, 17 isolates were uniquely identified in MGT, 15 isolates were uniquely identified by SNPs and 197 isolates were identified by both. The overall similarity in strain assignment to outbreak clusters as evaluated by kohens kappa was 0.913 and overall, 13 of 14 MGT clusters were identified by SNP analysis and 11 of 11 SNP clusters were identified in MGT analysis.
